# New Role of Red Blood Cells in Absorption of DNA Bearing Tumorigenic Mutations from Lung Cancer Tissue

**DOI:** 10.1101/2021.01.15.21249747

**Authors:** Nai-Xin Liang, Tao Wang, Cong Zhang, Zichen Jiao, Tianqiang Song, Hongwei Liang, Qihan Chen

**Author notes:** These authors share the first author.

## Abstract

Red blood cells (RBC) are commonly assumed to be vehicles for oxygen, carbon dioxide, and cells’ metabolic byproducts. In this study, we investigated whether RBC may contain cancer-cell derived DNA and whether such cargo may be used as a biomarker for detecting cancer. Using an *in vitro* co-culture system, we showed that RBC could absorb DNA bearing tumorigenic mutations from cancer cell lines. Next, we demonstrated that we could detect common genetic mutations, including *EGFR* 19 deletion, L858R, and *KRAS* G12 in RBC collected from early-stage non-small cell lung cancer patients. We were able to repeat our finding using both next-generation sequencing and droplet digital PCR. Our study highlights a new biological phenomenon involving RBC and their translational potential as a novel liquid biopsy technology platform that can be used for early cancer screening.

---

Red blood cells (RBC) are thought to have rich material exchange with cancer cells exhibiting very active metabolism (Fig. 1a). On the one hand, red blood cells provide oxygen and nutrients for the growth and development of cancer cells; on the other hand, red blood cells also rapidly take away carbon dioxide and metabolic byproducts produced by cancer cells. These lead to what tumor-derived cargo can be found in RBC and whether these molecules can be used to discriminate between cancer and healthy patients. It is well known that tumor cells release nucleic acids and proteins via exosomes, and these have been investigated as potential sources of cancer diagnostic biomarkers in liquid biopsy. RBC are found in much higher abundance than exosomes, and are known to interact directly with cancer cells and exosomes [1] and could adsorb nucleic acid directly from their environment [2]. RBC may take up tumor-derived nucleic acid through these interactions.

**Figure 1.**
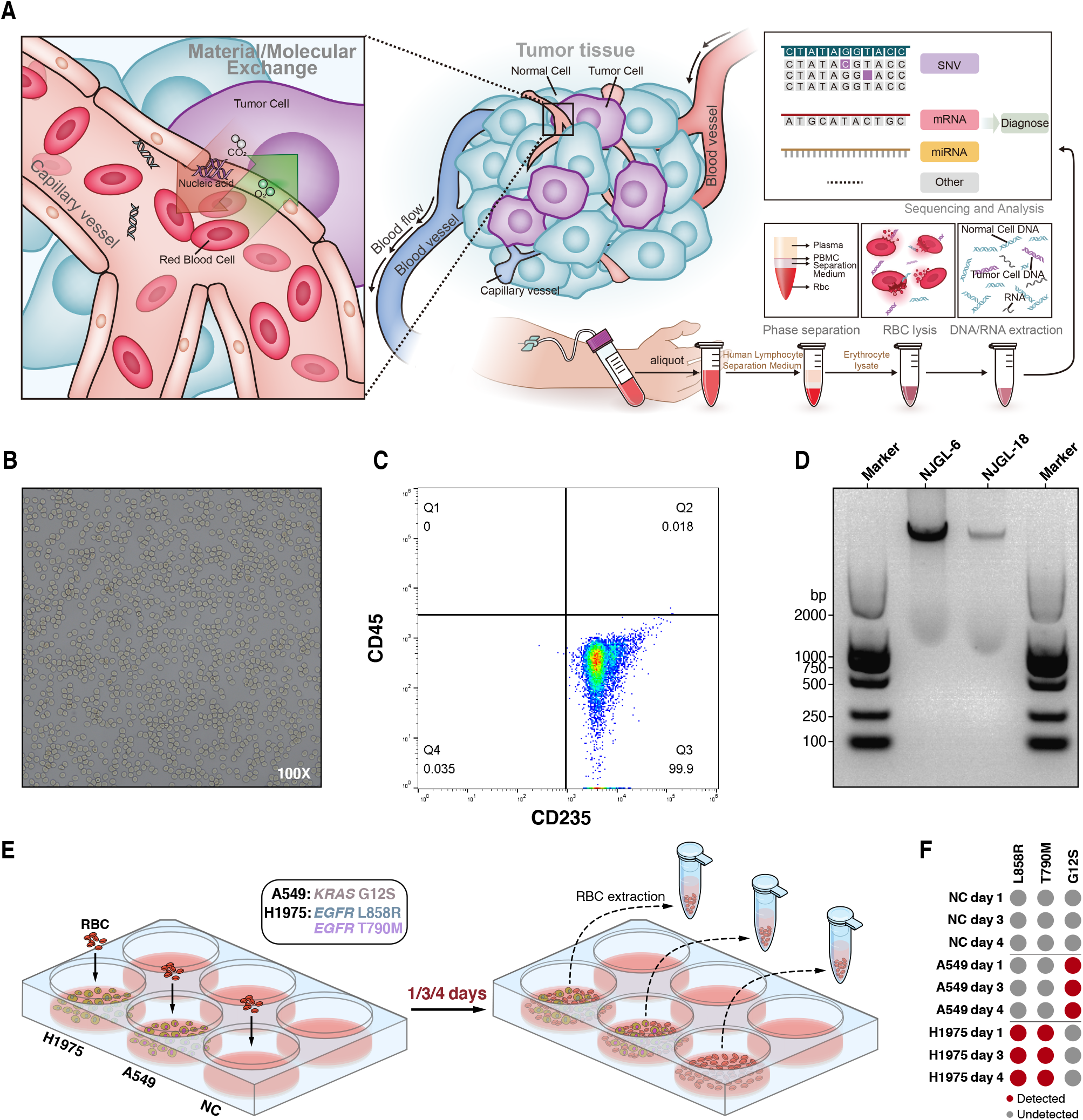
A. Schematic diagram depicting the exchange of cargo between tumor cells and red blood cells. RBC containing tumor cell-derived cargo may be used as a potential biomarker source for liquid biopsy cancer diagnostic. B. Light microscopy analysis of isolated red blood cells (100x magnification). C. Flow cytometry analysis detecting the presence of CD45 and CD235 to determine the purity of RBC preparation. D. Fragment sizes of DNA isolated from RBC determined by agarose gel. E. Schematic diagram depicting the *in vitro* co-culture system using RBC and cancer cell lines A549 and H1975. F. Detection of *EGFR* L858R, T790M and *KRAS* G12S in isolated RBC that was co-cultured with cancer cell lines.

We first speculated that cancer cells might have material exchange with RBC at an early stage of solid tumor development due to angiogenesis (Fig. 1A). To prove this hypothesis, we used a density gradient method to separate RBC from whole blood samples. We then determined the purity of our RBC preparation to ensure that subsequent analysis will not be contaminated by cargo found in other cell types. To confirm this, we examined the purity of our RBC preparation using both light microscopy at 100x magnification (Fig. 1B) and flow cytometry analyses with CD235a antibody for red blood cell and CD45 antibody for myeloid-derived nucleated cells in the blood (Fig. 1C).

Due to the lack of nuclei in mature RBC, it is assumed that these cells do not contain their own genomic DNA. Interestingly, DNA extracted from lung cancer patients demonstrated two main groups: larger than 2000 bps and between 1000 to 2000 bps (Fig. 1D). To test whether such DNA fragments could be transferred from other cells, we co-cultured purified RBC with lung cancer cell lines (Fig. 1E). The same mutations can be detected in RBC DNA as relative co-cultured cell lines but not in the RBC cultured alone (Fig. 1F).

These results implied that in our *in vitro* co-culture system, cancer cell-derived DNA fragments are definitely transferred into RBC. To further explore this phenomenon’s physiological significance, we carried out a clinical study to verify the substance exchange between cancer cells and RBC by targeting mutations produced in cancer cells but not in normal cells.

We analyzed the tissue and RBC samples of 28 patients, including 18 cases from Nanjing Drum Tower Hospital and 10 cases from Beijing Union Medical College Hospital. In terms of lung cancer staging, there were 21 patients in the first stage, accounting for 72% of the patient pool, containing 4 IA1, 8 IA2, 3 IA3, and 6 IB cases. Therefore, our sample composition is mainly composed of very early lung cancer patients. Notably, due to NSCLC surgical patients’ particularity in Union Medical College Hospital, we encountered 5 patients with multiple primary lung cancer tumor loci, which increased the difficulty of accurate detection.

Based on the NGS analysis and using tissue mutation status as a reference, a total of 6 *EGFR* 19del, 8 *EGFR* L858R, 2 *KRAS* G12 were all detected in DNA extracted from RBC and classified as true-positive results (Fig. 2A, Table 1). The IHC results of pathology or the NGS results of the third party were used as the reference. However, we detected 5 “false-positive” results in both *EGFR* 19del and L858R and 3 “false-positive” results in *KRAS* G12 detection, which we will elaborate on in the following part. In these 13 “false-positive” results, we found that in 4 patients, our tissue NGS test results were inconsistent with IHC or third-party tissue NGS results.For example, the tissues we obtained for samples No.9 and No.15 were L858R negative in NGS analysis, but tissue IHC and RBC NGS analysis from the same patients showed L858R positive (Table 1). For patient No.16, our tissue NGS of 19del was negative, but third party tissue NGS of different loci and RBC NGS was positive. Three other “false-positive” patients did not have comparable IHC results or third-party NGS results, so we could not evaluate whether their “false-positive” status was caused by tissue sampling issues or truly “false-positive”.

**Figure 2.**
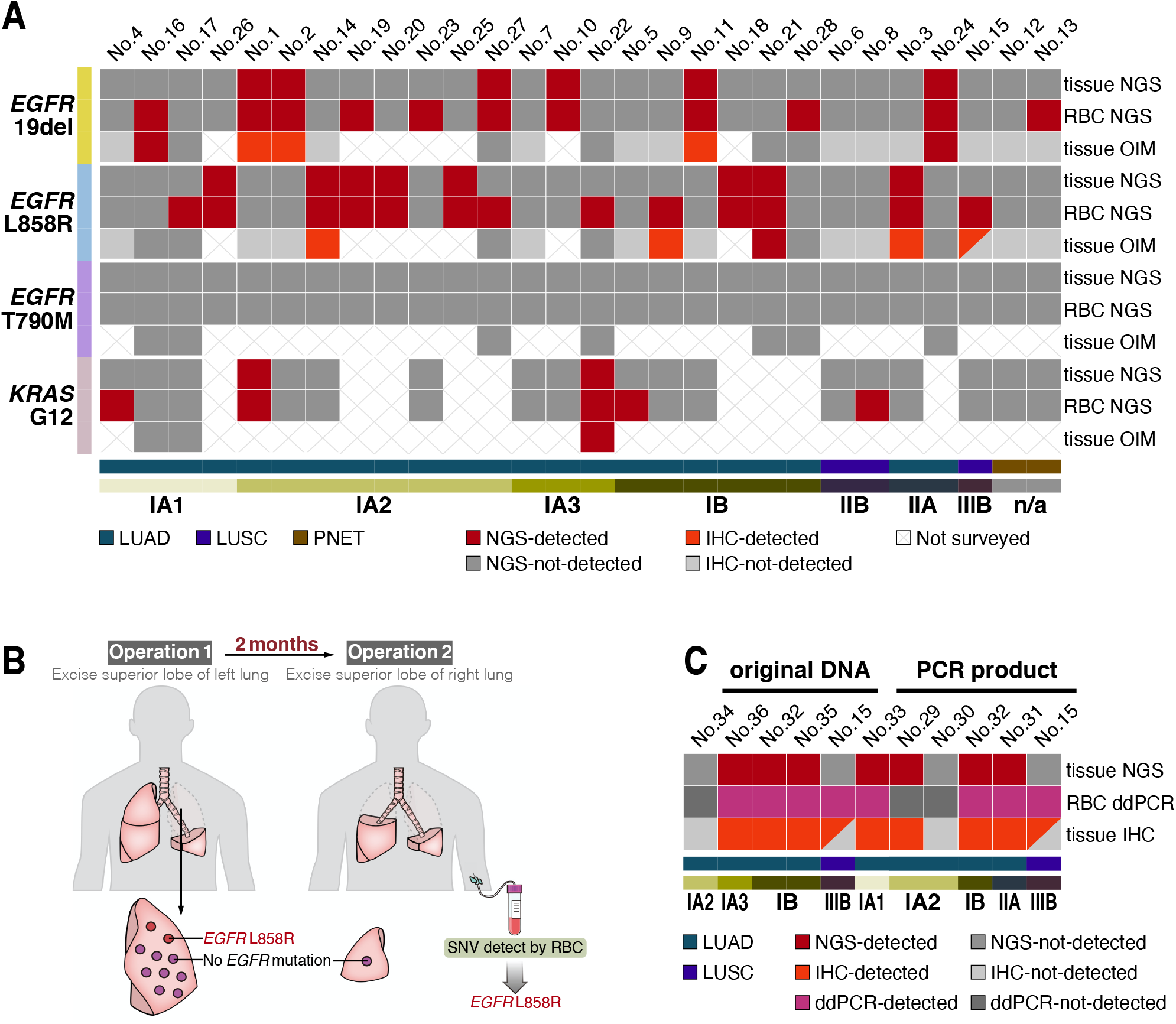
A. Heat map showing the detection of *EGFR* 19del/L858R/T790M and *KRAS* G12 in patient tissue or red blood cells. The methods used to detect the mutations include next-generation sequencing (NGS) or other independent measures (OIM) such as immunohistochemistry. Sample IDs are labeled at the top of the panel. B. Schematic diagram depicting the surgical process and sample collection steps for patient No.27. C. Heat map showing the detection of *EGFR* L858R in patient tissue using NGS or IHC and RBC using ddPCR. Both original DNA directly obtained after extraction, and PCR product obtained after the amplification of extracted DNA, were used in our analysis. Sample IDs are labeled at the top of the panel.

**Table 1.**
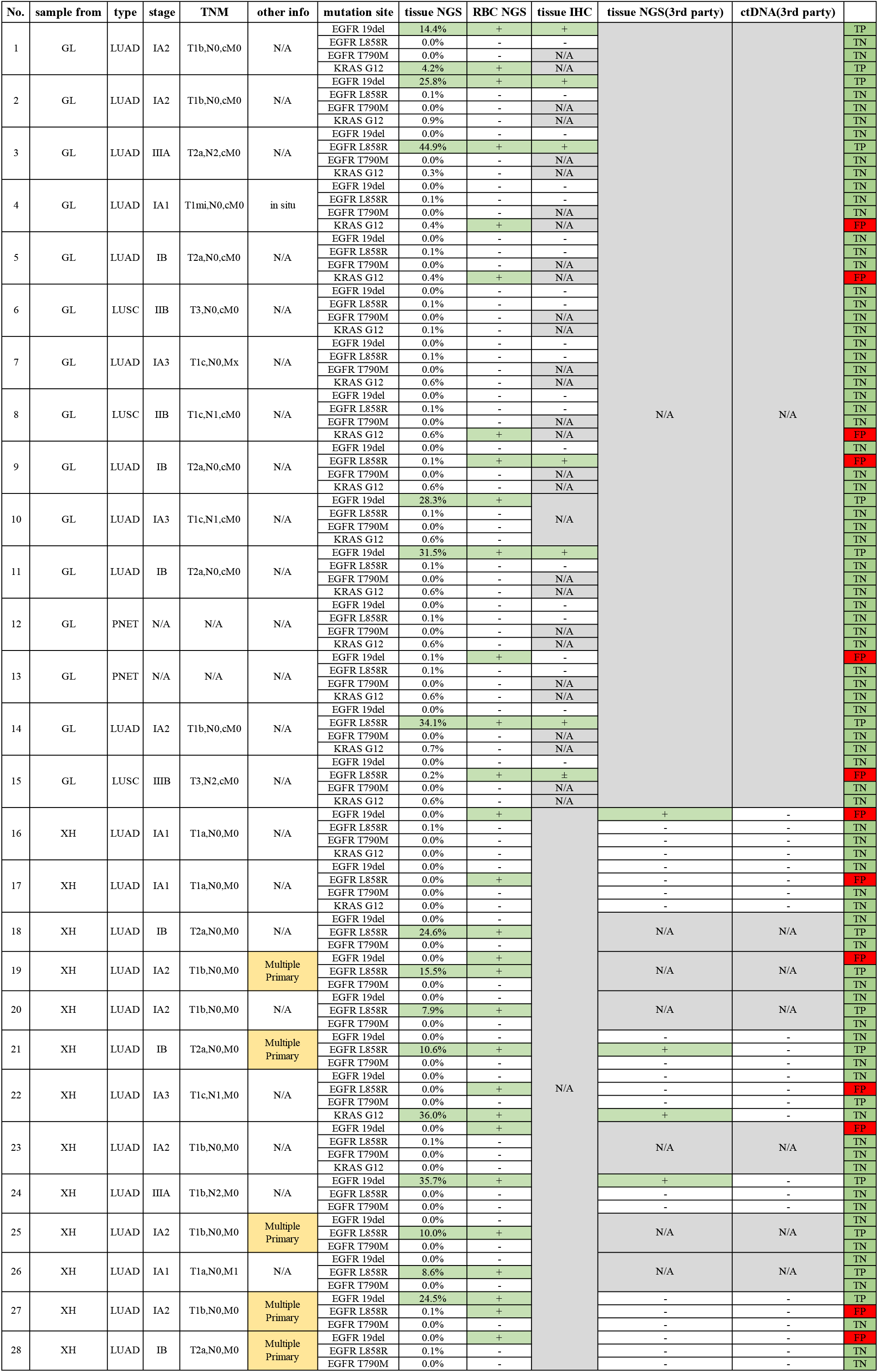

Interestingly, patient No.27, who had multiple primary lung adenocarcinomas and nodules on separate lobes, underwent two consecutive operations in a short period (Fig. 2B). The patient underwent the first operation to remove the left lung superior lobe that contained 9 cancer nodules. After two months, the patient underwent a second operation to remove the right lung superior nodule with one cancer nodule. In our study, we were first given obtained tissue samples and RBC samples from the second operation. Our results showed no *EGFR* L858R mutation in the tissue, but L858R mutation was detected in RBC. Fortunately, tissue samples from both operations were sent to a third-party for NGS. Although no L858R mutation was detected in the cancer tissue from the second operation, L858R mutation was detected in the first operation’s two largest nodules. These data revealed the occurrence of tumor tissue heterogeneity in this patient while also revealing that cancer cell-derived DNA fragments may remain in patients’ red blood cells for at least two months, which is quite different from the biology of cfDNA. Other multiple primary cancer patients with “false-positive” status, sample No.19 and No.28, may have the same issue. However, we were not able to obtain tissues from additional nodules to further verify these claims.

Since droplet digital PCR has a better sensitivity to detect low-frequency mutations, we used ddPCR further analyze 9 additional patient samples from Drum Tower Hospital to confirm the existence of cancer cell-derived DNA fragments carrying *EGFR* L858R mutations in RBC (Fig. 2C; Table 2). The samples were divided into two variations for analysis: 6 samples directly used DNA extracted from RBC as a template, and 6 samples used DNA as a template for PCR amplification first and then using the purified products as a template. Sample No.15 and No.32 were replicated in both variations of experiments. It can be seen that the RBC L858R results measured by ddPCR are generally consistent with the corresponding tissue L858R mutation detection by IHC, except a false negative result of sample No.29 that used its PCR amplified product for ddPCR analysis. Again, *EGFR* L858R were detected in both extracted DNA and amplification products in sample No.15 as NGS results. Once again, these data proved that RBC contained DNA fragments from cancer cells through material exchange, and was not caused by PCR amplification artifacts.

**Table 2.**
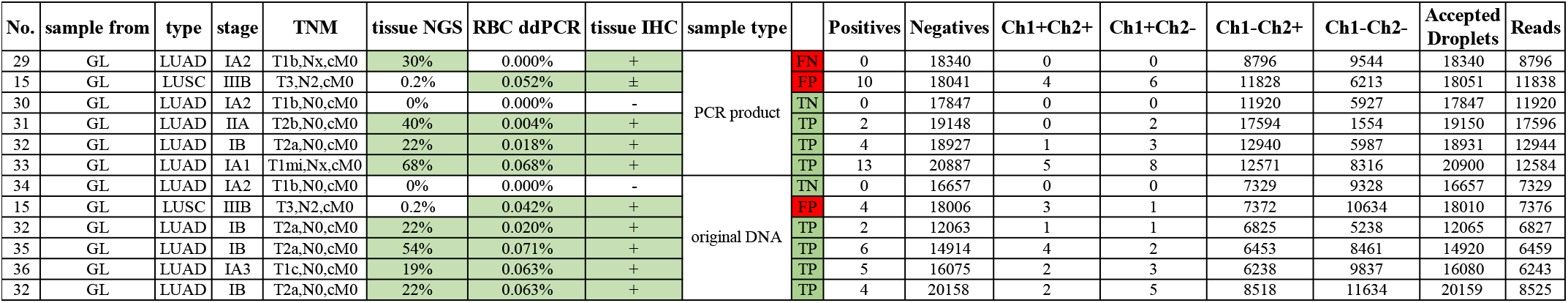

## Discussion

Mature red blood cells no longer have full nuclear and their own genomic DNA, but whether they have externally-derived DNA as cargo and whether they can be further transcribed and translated have been alluded to in previous studies[3, 4]. In addition, as the principal cells of oxygen and nutrient transport, red blood cells play an essential role in the occurrence and development of cancer[5]. This study explored whether RBC absorbed cancer cell-derived DNA, and whether this cargo can be used as a biomarker for early-stage cancer. Through *in vitro* studies, we showed that RBC can absorb DNA fragments from cancer cell lines and that these fragments carry specific mutations characteristic of these cells. We further demonstrated that this phenomenon exists under physiological conditions by showing that cancer cell-specific DNA can be detected in RBC collected from NSCLC patients, even at the earliest stages. Overall, our study indicates that molecular exchange between RBC and tumor cells is more complicated than previously understood. The cargo found within RBC may play a role in cancer occurrence and development and can be exploited as a biomarker for early cancer screening.

An important question in understanding our finding’s biological significance is the nature of DNA and the mechanism by which they are transferred into RBC. While a live cell cannot separate a piece of genomic DNA and give it to another cell while it is alive, cancer cells are known to produce circular extra-chromosomal DNA (ecDNA) to drive tumor pathogenesis and evolution [6]. ecDNA release from the cancer cell can be achieved through different mechanisms, including large extracellular vesicles, exosomes, or extracellular particles like exomeres or chromatimeres [7]. At this point, we speculate that RBC may be able to absorb these particles, consequently taking up the associated cancer cell-derived ecDNA. We are currently conducting follow-up studies to investigate the potential mechanisms of this phenomenon.

Our study also highlights the potential translational application of our discovery. Specifically, we demonstrated that we were able to consistently detect common *EGFR* and *KRAS* mutations using RBC collected from early-stage NSCLC patients, implying that our discovery can form the basis of a new liquid biopsy technology. One of the issues in our study was that we seemed to obtain a high false-positive rate. However, we also showed several examples that the false-positive results were due to inaccurate detection of tissue mutations due to tumor heterogeneity. In our follow-up study, we will optimize our tissue sampling method, extract DNA from multiple parts of cancer tissues and cover more primary nodules to obtain accurate tissue mutation results to the greatest extent. Another possible reason is the introduction of random mutations in multiple rounds of PCR and NGS sequencing, which may not be ignored in the detection of single point mutations. To eliminate this risk, multiple sample repeats will be introduced in the future study. Furthermore, the ddPCR results suggested that when the proportion of mutation fragments in the sample is deficient, the PCR process may further reduce the proportion of mutation fragments due to the sampling deviation, resulting in the inaccurate of the final detection results. Therefore, the optimization of PCR sampling deviation and enrichment of mutation fragments should be considered.

In conclusion, the use of RBC as a new liquid biopsy platform can greatly advance the stage of detection of NSCLC patients, thereby significantly improving his/her treatment options and survival rate. Furthermore, because the isolation and DNA extraction steps with RBC are much more manageable than cfDNA, mutation information can be obtained at a very low cost by simple amplification and NGS process. We believe that additional cancer-specific cargo can be found in RBC since we can detect cancer cell-derived DNA fragments in very early cancer patients,. For example, the specific epigenetic modification site of DNA[8, 9], the specific mRNA transcripts[10, 11], the specially modified protein of cancer cell[12, 13], specific glycosylation modification of lipids [14]. Therefore, RBC-based liquid biopsy represents a promising new tool for early cancer detection that warrants further investigation.

## Method

### 1. sample collection

This study recruited 50 patients with suspected lung cancer in thoracic surgery. The specific inclusion and exclusion criteria are shown in the supplementary table. The blood samples were collected within 1 h before the biopsy, 1 h before or during operation, and were further separated into plasma, PBMC, and RBC based on the description in the method part; the tissue samples were puncture samples or surgical tissue samples, which were further diagnosed as lung cancer by the hospital pathology department through HE staining, and then the cancer tissues in the whole tissue samples were further separated based on the pathological sections. Based on the pathology results, 42 samples from patients were finally applied in the study.

The study was divided into two centers, Peking Union Medical College Hospital and Nanjing Drum Tower Hospital. Based on the different hospital situations, the research plans of the two centers were slightly different: in Peking Union Medical College Hospital, we completed the separation of blood samples locally in Beijing and transported the separated RBC and tissue to the laboratory in Nanjing within 7 days to complete the subsequent NGS or ddPCR test; Peking Union Medical College Hospital screened the diagnosis of the same patient based on NGS in a third-party company as well as the mutation detection based on serum cfDNA. In Nanjing Drum Tower Hospital, we completed the separation of blood samples and the following NGS or ddPCR in Nanjing laboratory, and we screened mutation of cancer tissue through NGS; the pathology department completed the detection of L858R and 19del mutations of *EGFR* of the same patient by tissue IHC staining method. Follow up statistics based on our RBC detection results and tissue detection results for statistical comparison. The tissue IHC or third-party NGS results were considered as a reference. The results of the first batch of XX patients were disclosed. The relevant research has been approved by the ethics committee of the two hospitals.

### 2. RBC isolation

Whole blood was taken from patients and collected in Vacutainer tubes containing EDTA. Blood samples were processed within 12 h of collection. Separation of RBC using the density gradient method. First, the blood was separated by centrifugation (Eppendorf, Centrifuge 5810R) at 150 g for 10 min, and the supernatant was removed. Then, add an equal volume of PBS to the precipitate and mix gently. The diluted precipitate was then slowly added dropwise to 1.5 times the Human Lymphocyte Separation Medium’s precipitate volume and centrifuged at 800 g for 15 min, then slowly remove the supernatant. Third, add an equal volume of PBS to the precipitate, mix gently, and then centrifuged at 150 g for 15 min. The supernatant was removed, and RBC was then stored at −80 °C until DNA extraction.

### 3. Flow Cytometry

Red blood cells were incubated with FITC anti-human CD45 Antibody (HI30) (Biolegend, USA) and PE anti-human CD235a (Glycophorin A) Antibody (HI264) (Biolegend, USA) for 60 min. Subsequently, the red blood cells were washed three times with PBS for 5 min each in the dark and analyzed by a Becton Dickinson FACScan.

### 4. RBC co-culture with H1975 and A549 cells

H1975 and A549 cells were seeded at a density of 1□×□104 cells per well in the 24-well plates. After 24 hours, the obtained red blood cells, which were isolated from healthy donors by using Ficoll-Paque density centrifugation) were added to each well at a ratio of 0.5:1. Red blood cells were collected from the co-culture system on day 1, 3 and 4, and purified by Ficoll-Paque density centrifugation. The purification of red blood cells was confirmed by microscope and Flow Cytometry.

### 5. Amplicon NGS sequencing

According to the manufacturers ‘instruction, DNA from RBC and tumor tissues was extracted using TIANamp Genomic DNA Kit (Tiangen). PCR reactions were prepared in 50-μl volumes containing 1× Ex Taq Buffer (Mg2+ plus) (Takara), 0.4 μM of each primer, 0.2 mM of each of the four deoxynucleoside triphosphates (dNTPs), 10-100ng of template DNA, 1.25 U of TaKaRa Ex Taq HS (Takara). The PCR conditions consisted of a denaturation period at 98□ for 2 min followed by 35-38 cycles at 98□ for 10 s, 60□for 30 s, and 72□ for 30 s, followed by a final elongation at 72□ for 5 min. The PCR products were purified using the AxyPrep PCR Clean-Up Kit (Axygen) according to the manufacturer’s instructions and quantified by Nanodrop One (Thermofisher). Purified PCR products were used for library construction, and sequencing was performed in paired-end 150 mode with Illumina HiSeq X Ten.

### 6. Mutation detection by ddPCR

Bio-Rad QX200 did both original DNA and amplification products ddPCR with EFGR L858R mutation detection kit following the manual. Amplification products were prepared by a two-step PCR: the first-round PCR reaction was done with only forward primer for 15 cycles in 50-μl volumes containing 1×TransStart FastPfu Buffer (Transgen), 0.2 μM forward primer, 0.2 mM of each of the four deoxynucleoside triphosphates (dNTPs), 5-10ng of template DNA, 2.5 units of FastPfu DNA Polymerase (Transgen). For the second-round PCR, 0.2 μM reverse primer was added to the reaction solution to amplify for 25 cycles. The PCR products were purified using AxyPrep PCR Clean-Up Kit (Axygen) according to the manufacturer’s instructions and quantified by Nanodrop One (Thermofisher).

## Data Availability

all relevant data is shown in the tables provided.

